# Navigating transition: A mixed methods study protocol for understanding chronic pain, resilience, and well-being among Canadian Armed Forces Veterans, families, and service providers

**DOI:** 10.1101/2025.10.02.25337191

**Authors:** Jenny JW Liu, Erin Collins, Natalie Ein, Julia Gervasio, J. Don Richardson

## Abstract

**Introduction:** Transitioning from military to civilian life is often challenging for Canadian Armed Forces (CAF) Veterans, and chronic pain further complicates reintegration and wellbeing. Family members and service providers play critical but understudied roles in this process.

**Methods:** This mixed-methods study combines secondary analysis of the previous dataset with new qualitative interviews. Quantitative analyses will describe characteristics of Veterans with chronic pain and assess associations between pain severity and transition outcomes using regression models. Qualitative interviews with Veterans (n=10), family members (n=10), and service providers (n=10) will explore pain management, barriers and facilitators to care, and resilience strategies, analyzed using both deductive (framework-guided) and inductive thematic approaches.

**Results:** Findings will identify correlates of chronic pain and transition outcomes, as well as systemic gaps and resilience-enabling supports across the transition continuum.

**Discussion:** This study is the first in Canada to examine chronic pain in Veterans’ transition through a multi-system lens inclusive of family and provider perspectives. Grounded in resilience frameworks, the design addresses individual, relational, and structural influences on post-service adjustment. Anticipated findings will inform targeted interventions, caregiver supports, and provider education to strengthen resilience and improve care coordination for Veterans with chronic pain.

## Introduction

The transition to life after service is one of the most difficult and complex challenges reported by Veterans. Approximately 1 in 4 Canadian Armed Forces (CAF) Veterans report a difficult transition marked with mental health concerns and/or chronic pain (1–3). Difficulties encountered during this transition often contribute to, and may exacerbate, poor mental and physical health outcomes. A recent epidemiological study on chronic pain among Canadian Veterans found that 64% experienced either constant or intermittent chronic pain, while 25% experienced pain that significantly interfered with activities of daily living (2). For those navigating transition-related stressors, chronic pain can limit the capacity to take on new roles and duties and amplify existing challenges and barriers to successful reintegration.

Chronic pain and discomfort among Veterans have been linked with diminished quality of life (4) and an increased risk of suicide [e.g., (5,6)]. Moreover, Mikolas et al. (7) highlighted a shortage of services equipped to support the complex, multidimensional needs of Veterans. These findings underscore a need for a more comprehensive understanding of the impact of transition on Veterans’ access to, and uptake of, supports and services. They also indicate opportunities to expand and tailor interventions aimed at fostering resilience and improving overall wellbeing among Veterans.

Critically, the roles of service providers, family members, and close friends are increasingly recognized as integral to Veterans’ transition experiences. Service providers such as primary care physicians, pain specialists, allied health professionals, case managers, and service agents play a pivotal role in shaping treatment pathways and long-term health outcomes. Their insights into system-level barriers, resource constraints, and patient-provider misalignments offer important context to understanding the broader landscape of chronic pain care during the post-service transition. Also vital, family members and close friends often take on caregiving responsibilities—frequently without adequate preparation or support (8,9). Their involvement not only buffers Veterans against isolation but also contributes directly to care coordination, emotional regulation, and daily functioning. However, assuming these roles can come at a cost; families supporting a Veteran with chronic pain may experience significant emotional and psychological strain (10). Despite their significance, the perspectives and experiences of family members remain markedly understudied in the Canadian context, particularly in relation to their influence on the transition process.

### Theoretical Frameworks

This study employs two complementary frameworks: the Veteran Well-being Surveillance Framework and the Multi-System Model of Resilience (MSMR). Both frameworks contribute uniquely to a comprehensive understanding of the complex, multidimensional nature of chronic pain during the transition from military to civilian life.

The Veteran Well-being Surveillance Framework conceptualizes resilience as a multidimensional construct encompassing seven domains: purpose, finances, social integration, life skills, housing and physical environment, cultural and social environment, and health (11). This framework highlights how these domains collectively shape the wellbeing of Veterans and identifies priority areas that may be affected during and after transition.

In parallel, the MSMR (12,13) defines resilience as a dynamic process of adapting to challenges through leveraging internal and external resources. The MSMR adopts a bi-factor model that spans nine resource systems: health and wellness, life satisfaction, social security, health reserve, growth capacity, societal functioning, psychological regulation, self-esteem, and accessible supports. By attending to both individual capacities and structural/environmental influences, the MSMR is particularly well-suited to contextualize the evolving and intersecting challenges faced by Veterans, service providers, and families.

Together, these frameworks provide an integrated lens to explore how chronic pain disrupts various systems of functioning, and how resilience may be supported across multiple processes and systems. Importantly, both frameworks account for social and structural influences, and thus align with the study’s goal of understanding the roles of service providers and family members not as peripheral, but as integral to shaping transition trajectories. This dual-theory approach underscores the need to address Canadian-specific service gaps, cultural factors, and policy contexts in designing effective, tailored supports.

### Research Aims

Despite growing recognition of the impacts of chronic pain on Veterans’ quality of life, little is known about how these experiences are shaped by the broader military-to-civilian transition environment, including the perspectives of service providers and family members. This research addresses a significant gap in the literature by explicitly examining the intersection of chronic pain and transition, while integrating the voices of those most intimately involved in Veterans’ support networks. Moreover, Veterans, service providers, and family members are uniquely positioned to identify enduring gaps in the availability, accessibility, and effectiveness of services and supports for Veterans living with chronic pain, and to recommend opportunities for improvement across the transition continuum.

This paper outlines the protocol for a study designed to examine the lived experiences and systemic challenges faced by Canadian Armed Forces (CAF) Veterans with chronic pain, as well as their family members and service providers. The study aims to capture individual and structural factors shaping these experiences, and to detect gaps and opportunities within the existing support infrastructure. Employing a sequential mixed-methods design, the research integrates quantitative survey data with qualitative insights from semi-structured interviews to address the following objectives:

1. Describe the characteristics and post-service outcomes of Veterans with chronic pain.
2. Assess the relationship between chronic pain and transition experiences.
3. Evaluate the relevance of the MSMR and the Veteran Well-being Surveillance Framework in capturing key themes related to transition, chronic pain, resilience, and well-being.
4. Identify major barriers and facilitators in accessing support systems, including health services, case management, and informal care.
5. Develop recommendations for policy and practice to improve the transition experience, acknowledging the interconnected roles of Veterans, providers, and families.
6. Ultimately, this study seeks to inform a more nuanced and systemic understanding of chronic pain in transitioning Veterans—one that recognizes the interplay between individual experiences and the broader social and institutional contexts in which they unfold.

## Methods

### Overall Study Design

This study will explore resilience capacity, individual and systemic strengths, resource accessibility and effectiveness, and service utilization among CAF Veterans with chronic pain through a combination of self-reported surveys and in-depth interviews. A comprehensive needs assessment will be conducted to identify unmet needs across the domains of the Veteran Well-being Surveillance Framework, while also evaluating this framework’s alignment with the MSMR. Drawing on the perspectives of Veterans, family members, and service providers, the study will identify services and supports perceived to be effective, absent and needed, or available but presenting significant barriers to access and utilization. This approach will yield a holistic understanding of the systemic gaps and opportunities for enhancing care and support for Veterans with chronic pain.

The protocols of this study were reviewed and approved by the research ethics board at Western University (WREM #124958) and Lawson Research Institute (REDA #14617). <u>Recruitment and data collection are currently in progress, with recruitment anticipated to conclude by November 2025 and data collection and cleaning by January 2026. Final results are expected by May 2026.</u> Employing a mixed-methods approach, this study will analyze quantitative and qualitative data in parallel to develop a comprehensive understanding of the experiences of CAF Veterans living with chronic pain, as well as their family members, and service providers. Table 1 summarizes how each objective will be addressed using quantitative or qualitative methods. Detailed methodologies for each component are outlined in subsequent sections.

**Table 1.** Objectives by Analytical Approach and Phase of Military-to-Civilian Transition.

|  |  |  |
| --- | --- | --- |
| **Objective** | **Analytical approach** | **Phase of transition** |
| 1. Describe the characteristics and post-service outcomes of Veterans with chronic pain. | Quantitative: Descriptive statistics on demographics, service-related factors, release terms, and post-service supports, well-being, and resilience stratified by pain status. | Post-transition |
| 2. Assess the relationship between chronic pain and transition experiences. | Quantitative: Linear regression analysis to examine the relationship between chronic pain and transition scores, incorporating potential risk factors and mediators. | Post-transition |
| 3. Evaluate the relevance of the Multi-System Model of Resilience and the Veteran Well-being Surveillance | Qualitative: Deductive analysis of post-service experiences, categorized within | During and post-transition |
| Framework in capturing key themes related to transition, chronic pain, resilience, and well-being. | predefined domains using established frameworks. |  |
| 4. Identify major barriers and facilitators in accessing support systems, including health services, case management, and informal care. | Qualitative: Inductive analysis to determine barriers, facilitators, and opportunities for improvement across the transition continuum. | Pre-, during, and post-transition |
| 5. Develop recommendations for policy and practice to improve the transition experience, acknowledging the interconnected roles of Veterans, providers, and families. |  |  |

### Quantitative analysis

Objectives 1 and 2 will be addressed using data from a 2023 INSPIRE dataset, which contains survey responses from Canadian Veterans. Descriptive statistics will be generated to summarize respondent characteristics and outcomes, including demographics, service-related factors, terms of release, post-release supports, well-being, quality of life, and resilience. Respondents will be stratified by pain status, comparing those reporting moderate-to-severe pain interference with daily activities to those without. For continuous variables, independent t-tests will be used to assess differences between groups. In cases where normality assumptions are violated, Mann-Whitney U tests will be applied. For categorical variables, chi-square tests will be conducted, with Fisher’s exact tests used where cell counts are low. Effect sizes with 95% confidence intervals will be reported for all comparisons, including Cohen’s *d* for continuous outcomes and Cramér’s *V* (or the *phi* coefficient for 2-by-2 tables) for categorical outcomes.

Missing data will be examined for both extent and pattern. If fewer than 5% of observations are missing on a given variable and the pattern appears random, listwise deletion will be applied. Variables with more extensive or systematically patterned missingness will be addressed using multiple imputation by chained equations (MICE).

To evaluate the relationship between chronic pain and transition scores (measured using the Transition to Civilian Life Scale [TCLS]; α = .915), linear regression analysis will be performed. The model will adjust for covariates identified a priori, including age, education, length of service, marital status, sex/gender, months since release, number of deployments, service branch and class, terms of release, and household income. Additionally, the potential mediating effect of health-related impacts on social activities will be examined.

All quantitative analyses will be conducted using RStudio (version 4.4.1).

## Qualitative analysis

### Participant Selection and Recruitment

This study will engage three groups of participants: (1) CAF Veterans with chronic pain, (2) family members of CAF Veterans with chronic pain, and (3) service providers for CAF Veterans with chronic pain. Participants will be recruited through e-mail invitations sent to consent-to-contact lists from prior projects at the MacDonald Franklin Operational Stress Injury Research Centre (MFOSIRC), as well as through word-of-mouth referrals and targeted outreach via professional networks. We will seek to engage 30 participants total (10 CAF Veterans, 10 family members, and 10 service providers). All eligible participants must be English-speaking, aged 18 years or older and can include both male and female individuals. CAF Veterans must be Canadian Veterans living with chronic pain. Family members must be related to a CAF Veteran living with chronic pain. Service providers must be actively involved in supporting Veterans living with chronic pain, and include service agents, case managers, and physicians and allied health professionals.

### Interviews

Semi-structured interviews were developed for each participant group to probe experiences of chronic pain from these varying perspectives across 8 question categories: (1) general experience with Veteran supports, (2) chronic pain management, (3) impacts on daily life, (4) challenges and facilitators affecting the availability, accessibility, effectiveness of supports (5) well-being domains, (6) risk and resilience strategies, and (7) additional insights and closing reflections. Interview questions relating to impact on daily life and well-being domains will specifically probe the Veteran Well-Being Surveillance Framework domains and include questions on employment, purposeful activities, finances, physical and mental health, life skills, social integration, housing and physical environment, and cultural/social environment. Interviews were developed via collaboration between research experts and individuals with lived experiences (i.e., CAF Veterans). Examples of questions for each participant group can be found in Table 2.

**Table 2.** Examples of Interview Questions for Question Categories and Participant Groups.

| Question Categories | Participant Group |  |  |
| --- | --- | --- | --- |
|  | CAF Veterans | Family Members | Service Providers |
| *General Experience with Veteran Support* | What were the most challenging aspects of your transition? | What have been the most challenging aspects of supporting a Veteran during their transition? | What are the primary challenges you observe Veterans facing during their transition from military to civilian life? |
| *Chronic Pain Experiences / Management* | How long have you experienced chronic pain? | How does their chronic pain affect your role as a caregiver/supporter? | What types of treatments and services do you recommend or provide for Veterans experiencing chronic pain? |
| *Impact on Daily Life* | Does chronic pain disrupt any part of your life? If yes, how so? | How does the Veteran's chronic pain disrupt your daily life? | How does chronic pain affect Veterans' daily lives based on your observations and interactions? |
| *Challenges and Facilitators in Accessing Care* | Could you describe any challenges you have experienced while accessing care for chronic pain? | Can you describe any facilitators that have helped you and the Veteran access care for chronic pain more effectively? | What challenges do Veterans face when accessing care and support for chronic pain? |
| *Well-being Domains* | How does chronic pain influence your... [e.g., social integration]? | How does the Veteran's chronic pain influence your overall well-being in areas such as... [e.g., mental health]? | How does chronic pain influence the overall well-being of Veterans in areas such as... [e.g., physical health]? |
| *Risk and Resilience Strategies* | What strategies have you used to manage and live with chronic pain in your community? | What strategies do you use to manage your own well-being while supporting the Veteran? | What risk factors have you identified that affect Veterans' ability to cope with chronic pain? |
| *Additional Insights / Closing Thoughts* | What improvements or changes would you suggest to better support Veterans living with chronic pain? | Is there anything else you would like to add about supporting Veterans with chronic pain that we haven't covered? | Based on your experience, how do chronic pain issues shape Veterans' views on available support services? |

### Procedures

All potential participants (i.e., CAF Veterans, family members, and service providers) will first be provided with a Letter of Information and Consent via email, and can take as much time as they feel is needed prior to providing informed consent and proceeding with the study. Participants will be told that their participation is completely voluntary, and they can refuse to participate or withdraw from the study at any time. Each participant will receive a gift card as compensation for their participation, upon the completion of their interview. All interviews will be audio recorded for transcription purposes. Study links will remain active until the desired sample sizes are reached for each participant group.

### Group 1 - CAF Veterans with Chronic Pain

Veteran participants will be recruited from a subset of individuals who completed a prior needs assessment study focusing on their transition from military to civilian life. Eligible Veterans for the current study are those who previously indicated that chronic pain has moderately-to-severely affected their daily activities. These individuals will be contacted via email and invited to participate in a 1-hour virtual interview to share their experiences living with chronic pain and navigating the transition process, including accessing and utilizing available resources and services. Prior to the interview, participants will be asked to complete an optional 30-minute online survey assessing the nature and impact of their chronic pain using standardized measures, including the Multidimensional Scale of Independence (MSI) and Pain Self-Efficacy Questionnaire (PSEQ).

### Group 2 – Family Members

Family members who wish to participate in this study will be contacted via email and invited to participate in a 1-hour virtual interview regarding their experiences supporting a Veteran with chronic pain and navigating resources and services. Family members will also complete the PSEQ (with the interviewer) following the interview. This survey will take an additional 5 minutes to complete.

### Group 3 – Service Providers

Service providers who wish to participate in this study will be contacted via email and invited to participate in a 1-hour virtual interview regarding their experiences supporting a Veteran with chronic pain. No additional surveys will be completed by service providers.

### Characterization of Pain

Before the interview, CAF Veterans will complete a brief assessment of chronic pain, comprising two validated self-report pain measures. The usability and functionality of this survey were previously tested by the research team, including adaptive questioning, and branching functions.

#### Multidimensional Symptom Index [MSI; (14)]

The MSI is a 10-item patient reported outcome measure capturing both the frequency and degree of intensity of 10 somatic and non-somatic symptoms which contribute to the experience of living with pain. Symptom frequency is rated on a 4-point Likert scale ranging from 0 (Never) to 3 (Always). Symptom intensity is rated on a 4-point Likert scale ranging from 1 (Barely noticeable, doesn’t really bother me) to 4 (So intense I have to stop what I am doing and seek relief). In examining chronic pain in a cross-sectional context, Walton and Marsh (2018) note that the MSI can be used to identify high priority symptoms that have the greatest impact on pain experience and should be targeted for intervention.

#### Pain Self-Efficacy Questionnaire [PSEQ; (15)]

The PSEQ is a 10-item survey designed to quantify a person’s confidence in carrying out everyday activities despite persistent pain. Each item is rated on a 7-point rating scale from 0 (Not at all confident) to 6 (Completely confident). Item scores are summed to yield a total between 0 to 60, with higher scores indicating stronger pain self-efficacy. The PSEQ has been applied in Canadian Veteran cohorts (16) and aligns well with family-inclusive care, given items focus on everyday functioning, such as household tasks, work, and social engagement.

### Data Analytic Plan

Qualitative data from interviews will be analyzed using a combination of deductive and inductive reasoning. While a deductive approach offers several advantages, such as providing a structured analytical framework and facilitating comparisons with broader populations, it can lack the depth and contextual nuance necessary to fully capture the complexity of participants’ experiences. Through an inductive approach, we will allow themes to emerge organically, ensuring that interviewees’ perspectives are authentically represented.

An initial codebook will be developed to theory-informed and emergent themes. Interviews will be coded iteratively as data are collected; new themes will prompt updates to the codebook, and previously coded transcripts will be revisited as needed to ensure consistency. NVivo software will be used to support coding, theme identification, and qualitative synthesis. Analytic focus will be placed on identifying strengths, resources, and challenges related to chronic pain and transition, with attention to the evolving identities and lived experiences of each participant group. Select participant quotes will be included in reporting to preserve the authenticity and richness of individual narratives.

Deductive methods will be applied to address Objective 3. Responses related to post-service experiences will be systematically mapped onto the predefined domains of the Veteran Well-being Surveillance Framework (purpose, finances, social integration, life skills, housing and physical environment, culture and social environment, and health) and the levels of the MSMR (intra-individual, interpersonal, and socio-ecological factors). Subsequently, the capacity of each framework in capturing key themes related to transition, chronic pain, resilience, and well-being will be evaluated and compared.

Inductive methods will be employed to address Objectives 4 and 5. Qualitative interview data will be systematically analyzed to identify services and supports encountered across the military-to-civilian transition that participants perceive as: (a) effective; (b) needed but unavailable; and (c) available but ineffective. The analysis will consider both formal systems (e.g., health care, social services, case management) and informal supports (e.g., family networks, peer support). Using an inductive thematic analysis approach, patterns and themes will emerge directly from the data, offering insights into gaps, strengths, and barriers within existing support structures. Findings from the interviews will also inform the development of recommendations for enhancing care and support systems across the transition continuum. Particular attention will be given to identifying opportunities for meaningful and sustainable improvements that promote resilience and well-being. The results are intended to inform policy, guide program development, and contribute to the optimization of service delivery models for Veterans transitioning to civilian life.

Together, these qualitative analyses will complement the quantitative findings and ultimately strengthen the development of practical recommendations to improve support systems across the transition continuum for Veterans living with chronic pain.

## Discussion

This study protocol outlines a convergent mixed-methods investigation into the intersection of chronic pain and the military-to-civilian transition among CAF Veterans, with the explicit inclusion of family members and service providers. By incorporating both qualitative and quantitative methodologies, this design allows for a holistic exploration of how chronic pain interacts with post-service identity, service access, and overall wellbeing, while centering the lived realities of Veterans and their broader support networks.

A major strength of this study lies in its conceptual grounding in two robust frameworks: the Veteran Well-being Surveillance Framework and the MSMR. These models enable a multidimensional understanding of resilience that captures individual, relational, and structural influences on Veterans’ post-service adjustment. Moreover, the integration of family and provider perspectives adds a critical layer complexity to a field that has traditionally focused on the Veteran in isolation. Through the triangulation of these perspectives, we aim to generate insights that are not only descriptive but also actionable—shedding light on potential service gaps, unmet needs, and resilience-enabling supports.

Importantly, this study addresses a critical gap in Canadian research by prioritizing the voices of family members and service providers—groups whose contributions are vital but frequently underrecognized in both policy and empirical literature. For family members, caregiving demands can affect their own wellbeing, especially when they lack adequate training or systemic support. For service providers, navigating institutional constraints and managing patient-provider disconnects may limit their ability to offer effective chronic pain care. Capturing these realities through in-depth interviews offers a more complete picture of the systemic and interpersonal dynamics shaping the Veteran experience.

The anticipated findings will offer a valuable evidence base to inform policy and practice. Insights from this study can inform better co-designed interventions, including caregiver support programs, provider education in trauma-informed pain care, and service navigation tools tailored to transitioning Veterans. Ultimately, this study seeks not only to document experiences, but to serve as a catalyst for systems-level change—ensuring that Veterans living with chronic pain, and those who support them, are met with care systems that are responsive, integrated, and resilience-promoting.

## Data Availability

No datasets were generated or analysed during the current study. All relevant data from this study will be made available upon study completion.

## Acknowledgements

We would like to thank Robin Campbell and Cassandra Skrotzki for their valuable feedback and contributions to this manuscript. We would also like to thank Lisa Garland-Baird and the Veterans Affairs Canada (VAC) group for their support, insights, and collaboration, which greatly enhanced the quality and relevance of this work.

